# The Snoring Survivors

**DOI:** 10.1101/2023.02.20.23286207

**Authors:** Ikechukwu Ogbu, Bagrat Hakobyan, Christoph Sossou, Jeffrey Levisman, Chukwudi Obiagwu, Alfred Danielian

## Abstract

**Background:** The routine administration of supplemental oxygen to non-hypoxic patients with acute myocardial infarction has been abandoned for lack of mortality benefit. However, the benefits of continuous positive airway pressure (CPAP) use in patients hospitalized with acute cardiovascular disease and concomitant obstructive sleep apnea (OSA) remain to be elucidated.

**Methods:** Using ICD-10-CM codes, we searched the 2016-2019 Nationwide Inpatient Sample for patients diagnosed with unstable angina, acute myocardial infarction (AMI), acute decompensated heart failure (ADHF), and atrial fibrillation with rapid ventricular response (AFRVR), who also carried a diagnosis of OSA. We identified in-hospital CPAP use with ICD-10-PCS codes. In-hospital death, length of stay (LOS) and hospital charges were compared between patients with and without OSA, and between OSA patients with and without CPAP use.

**Results:** Our sample included 2,959,991 patients, of which 1.5% were diagnosed with UA, 30.3% with AMI, 37.5% with ADHF, and 45.8% with AF. OSA was present in 12.3%. Patients with OSA were more likely to be younger, male, smokers, obese, have chronic obstructive pulmonary disease, renal failure, and heart failure (*p* < 0.001 for all). Patients with OSA had significantly lower in-hospital mortality (aOR: 0.71, 95% CI [0.7-0.73]). Among patients with OSA, CPAP use significantly increased the odds of in-hospital death (aOR: 1.51, 95% CI [1.44-1.60]), LOS (adjusted mean difference of 1.49 days, 95% CI [1.43, 1.55]), and hospital charges (adjusted mean difference of $1168, 95% CI [273, 2062]).

**Conclusion:** Our study showed that patients with recognized OSA and hospitalized for AMI, ADHF or AFRVR, who were not treated with CPAP, had significantly lower in-hospital mortality and resource utilization.

## Introduction

Obstructive sleep apnea (OSA) manifests as recurrent partial or complete airway obstruction, resulting in chronic intermittent hypoxemia and autonomic dysfunction. The chronic intermittent hypoxia associated with OSA can result in hormonal and autonomic dysregulation, often manifesting as cardiac rhythm disturbances, such as atrial fibrillation, bradycardia and sudden cardiac death. Moreover, OSA is highly prevalent among patients hospitalized for acute cardiovascular disease, such as myocardial infarction, heart failure, atrial fibrillation and stroke^1^.

#### Clinical Perspective

##### What is New?

- This is the largest study examining the impact of CPAP treatment in sleep apnea patients hospitalized for myocardial infarction, heart failure, and atrial fibrillation.
- We showed that patients with sleep apnea who were treated with CPAP had higher in-hospital mortality, total hospital charges and longer stay duration.

##### What Are the Clinical Implications?

- Patients hospitalized for MI, HF or AF with known sleep apnea have decreased in-hospital mortality, but positive airway pressure utilization showed increased mortality.

Although several studies have observed a significant association between OSA and long-term adverse cardiovascular outcomes, there are fewer studies addressing the impact of treatment with continuous positive airway pressure (CPAP) among hospitalized patients with OSA and acute cardiovascular disease^2^. The largest randomized controlled trial (RCT) to date, the SAVE trial, randomized 2717 patients with established cardiovascular disease (a minority with HF) and moderate-to-severe OSA to CPAP plus usual care and to usual care alone. CPAP use did not reduce the composite cardiovascular endpoint, although in secondary analyses there was a lower risk of cerebrovascular events among patients using CPAP for at least 4 hours per night^3^. In another large single tertiary care center experience of roughly 94,000 ward admissions, OSA patients had lower odds of inpatient death (aOR = 0.70 [0.58-0.85]), transfer to ICU (aOR = 0.91 [0.84-0.99]), and cardiac arrest (OR = 0.72 [0.55-0.95]), all adjusted for important confounders^4^.

We analyzed the Healthcare Cost and Utilization Project Nationwide Inpatient Sample (HCUP NIS) to identify patients diagnosed with unstable angina (UA), acute myocardial infarction (AMI), acute decompensated heart failure (ADHF), and atrial fibrillation with rapid ventricular response (AFRVR), who had a concomitant diagnosis of OSA during their index hospitalization. We looked at CPAP utilization, in-hospital deaths, hospital length of stay (LOS), and total hospital charges among these patients.

## Methods

### Data Source

De-identified data from the 2016-2019 HCUP NIS^5^ was analyzed. The timeframe was chosen to avoid the myriad complex and confounding effects that the SARS CoV-19 pandemic could have potentially contributed in the years that followed. The HCUP NIS 2016-2019 dataset contains diagnoses and procedures encoded with 10^th^ International Classification of Diseases, Clinical Modification (ICD-10-CM) and Procedure Coding System (ICD-10-PCS) codes, respectively^6^. Additionally, several important clinical characteristics are further encoded using dataset-specific Agency for Healthcare Research and Quality (AHRQ) codes (e.g. diabetes or hypertension with or without complications, alcohol abuse, severe renal failure, etc.)^5,6^.

### Characteristics of the Study Sample

We identified age (categorized), gender, race, past or current tobacco use, and alcohol abuse for every inpatient record with the diagnoses of interest (UA, AMI, ADHF, AFRVR, OSA), along with the presence of important comorbidities (diabetes, hypertension, heart failure with reduced ejection fraction (HFrEF), heart failure with preserved ejection fraction (HFpEF), non-severe and non-end-stage, and severe or end-stage renal failure, obesity (not overweight), obesity hypoventilation syndrome (OHS; Pickwickian), central sleep apnea, chronic obstructive pulmonary disease (COPD), right ventricular hypertrophy (RVH) or cor pulmonale, type II pulmonary hypertension, type III pulmonary hypertension, any non-traumatic cerebrovascular accident (ischemic or hemorrhagic stroke, intracranial or intracerebral hemorrhage), sepsis (with and without shock), and inpatient treatment with CPAP. A full list of extracted items along with respective query codes can be found in **Supplemental Table 1**.

### Outcomes

Our primary outcome of interest was in-hospital death. Secondary outcomes included length of stay (LOS) and total hospital charges. We compared in-hospital death, LOS and total charges between patients with and without OSA, and between OSA patients who were and were not assigned to CPAP treatment during index hospitalization.

### Statistical Analysis

We ran a binary logistic regression analysis to determine the odds ratio of death during hospitalization (unadjusted) among patients with and without OSA, and for individual subgroups of UA, AMI, ADHF, AF, and patients with OSA who were and were not assigned to CPAP. To adjust for potential confounders, several covariates were introduced into the logistic regression model: sex, age category, race, smoking, alcohol abuse, obesity, hypertension, diabetes mellitus, any non-traumatic cerebrovascular accident, COPD, OHS, pulmonary hypertension (type II and III), *cor pulmonale* or RVH, HFpEF, HFrEF, end-stage (ESRD) or severe renal failure, non-end-stage chronic kidney disease (CKD) or non-severe renal failure, sepsis with and without shock. The means of LOS and costs were compared between subgroups of interest using one way ANOVA, and univariate analysis of covariates (ANCOVA) was used to adjust for confounders. A 2-tailed *p < 0*.*05* was used to denote statistical significance. All analyses were carried out using IBM SPSS v29.0 statistical software.

## Results

### Demographic and Clinical Characteristics

Our sample included 2,959,991 patients, of which 1.5% were diagnosed with UA, 30.3% with AMI, 37.5% with ADHF, and 45.8% with AFRVR. OSA was present in 12.3%. Patients with OSA were slightly younger, more likely to be male, obese, and past or current smokers (*p* < 0.001 for all). They were also more often comorbid with diabetes, renal failure, HFpEF, COPD, Cor Pulmonale or RVH, Pulmonary Hypertension, and treated with CPAP (*p* < 0.001). The complete demographic and clinical characteristics are shown in **Table 1**.

**Table 1.**
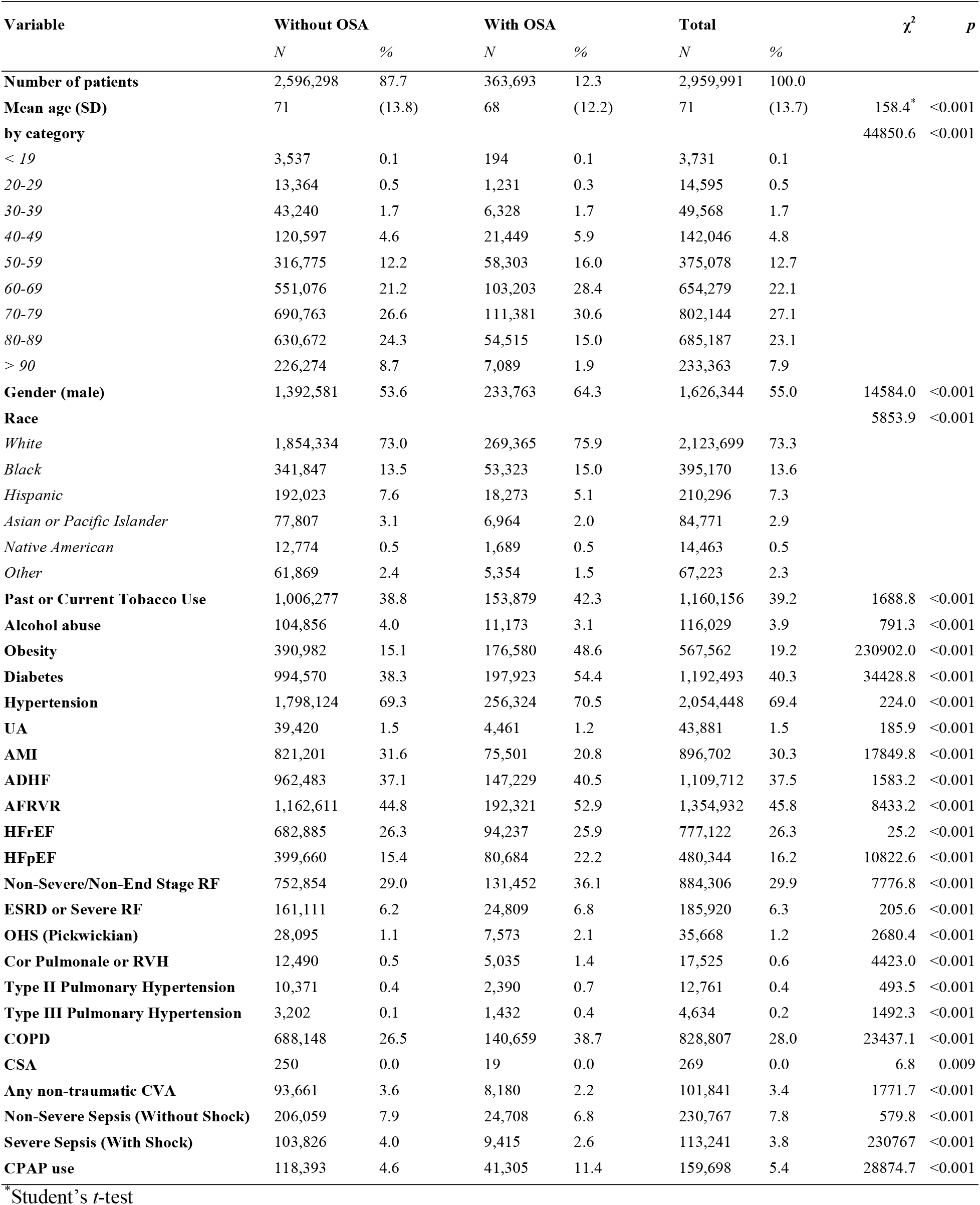
Demographic and Clinical Characteristics of the Study Population

| Variable | Without OSA | | With OSA | | Total | | $\chi^2$ | *p* |
| --- | --- | --- | --- | --- | --- | --- | --- | --- |
|  | *N* | % | *N* | % | *N* | % |  |  |
| **Number of patients** | 2,596,298 | 87.7 | 363,693 | 12.3 | 2,959,991 | 100.0 |  |  |
| **Mean age (SD)** | 71 | (13.8) | 68 | (12.2) | 71 | (13.7) | 158.4* | <0.001 |
| **by category** |  |  |  |  |  |  | 44850.6 | <0.001 |
| < 19 | 3,537 | 0.1 | 194 | 0.1 | 3,731 | 0.1 |  |  |
| 20-29 | 13,364 | 0.5 | 1,231 | 0.3 | 14,595 | 0.5 |  |  |
| 30-39 | 43,240 | 1.7 | 6,328 | 1.7 | 49,568 | 1.7 |  |  |
| 40-49 | 120,597 | 4.6 | 21,449 | 5.9 | 142,046 | 4.8 |  |  |
| 50-59 | 316,775 | 12.2 | 58,303 | 16.0 | 375,078 | 12.7 |  |  |
| 60-69 | 551,076 | 21.2 | 103,203 | 28.4 | 654,279 | 22.1 |  |  |
| 70-79 | 690,763 | 26.6 | 111,381 | 30.6 | 802,144 | 27.1 |  |  |
| 80-89 | 630,672 | 24.3 | 54,515 | 15.0 | 685,187 | 23.1 |  |  |
| > 90 | 226,274 | 8.7 | 7,089 | 1.9 | 233,363 | 7.9 |  |  |
| **Gender (male)** | 1,392,581 | 53.6 | 233,763 | 64.3 | 1,626,344 | 55.0 | 14584.0 | <0.001 |
| **Race** |  |  |  |  |  |  | 5853.9 | <0.001 |
| White | 1,854,334 | 73.0 | 269,365 | 75.9 | 2,123,699 | 73.3 |  |  |
| Black | 341,847 | 13.5 | 53,323 | 15.0 | 395,170 | 13.6 |  |  |
| Hispanic | 192,023 | 7.6 | 18,273 | 5.1 | 210,296 | 7.3 |  |  |
| Asian or Pacific Islander | 77,807 | 3.1 | 6,964 | 2.0 | 84,771 | 2.9 |  |  |
| Native American | 12,774 | 0.5 | 1,689 | 0.5 | 14,463 | 0.5 |  |  |
| Other | 61,869 | 2.4 | 5,354 | 1.5 | 67,223 | 2.3 |  |  |
| **Past or Current Tobacco Use** | 1,006,277 | 38.8 | 153,879 | 42.3 | 1,160,156 | 39.2 | 1688.8 | <0.001 |
| **Alcohol abuse** | 104,856 | 4.0 | 11,173 | 3.1 | 116,029 | 3.9 | 791.3 | <0.001 |
| **Obesity** | 390,982 | 15.1 | 176,580 | 48.6 | 567,562 | 19.2 | 230902.0 | <0.001 |
| **Diabetes** | 994,570 | 38.3 | 197,923 | 54.4 | 1,192,493 | 40.3 | 34428.8 | <0.001 |
| **Hypertension** | 1,798,124 | 69.3 | 256,324 | 70.5 | 2,054,448 | 69.4 | 224.0 | <0.001 |
| **UA** | 39,420 | 1.5 | 4,461 | 1.2 | 43,881 | 1.5 | 185.9 | <0.001 |
| **AMI** | 821,201 | 31.6 | 75,501 | 20.8 | 896,702 | 30.3 | 17849.8 | <0.001 |
| **ADHF** | 962,483 | 37.1 | 147,229 | 40.5 | 1,109,712 | 37.5 | 1583.2 | <0.001 |
| **AFRVR** | 1,162,611 | 44.8 | 192,321 | 52.9 | 1,354,932 | 45.8 | 8433.2 | <0.001 |
| **HFrEF** | 682,885 | 26.3 | 94,237 | 25.9 | 777,122 | 26.3 | 25.2 | <0.001 |
| **HFpEF** | 399,660 | 15.4 | 80,684 | 22.2 | 480,344 | 16.2 | 10822.6 | <0.001 |
| **Non-Severe/Non-End Stage RF** | 752,854 | 29.0 | 131,452 | 36.1 | 884,306 | 29.9 | 7776.8 | <0.001 |
| **ESRD or Severe RF** | 161,111 | 6.2 | 24,809 | 6.8 | 185,920 | 6.3 | 205.6 | <0.001 |
| **OHS (Pickwickian)** | 28,095 | 1.1 | 7,573 | 2.1 | 35,668 | 1.2 | 2680.4 | <0.001 |
| **Cor Pulmonale or RVH** | 12,490 | 0.5 | 5,035 | 1.4 | 17,525 | 0.6 | 4423.0 | <0.001 |
| **Type II Pulmonary Hypertension** | 10,371 | 0.4 | 2,390 | 0.7 | 12,761 | 0.4 | 493.5 | <0.001 |
| **Type III Pulmonary Hypertension** | 3,202 | 0.1 | 1,432 | 0.4 | 4,634 | 0.2 | 1492.3 | <0.001 |
| **COPD** | 688,148 | 26.5 | 140,659 | 38.7 | 828,807 | 28.0 | 23437.1 | <0.001 |
| **CSA** | 250 | 0.0 | 19 | 0.0 | 269 | 0.0 | 6.8 | 0.009 |
| **Any non-traumatic CVA** | 93,661 | 3.6 | 8,180 | 2.2 | 101,841 | 3.4 | 1771.7 | <0.001 |
| **Non-Severe Sepsis (Without Shock)** | 206,059 | 7.9 | 24,708 | 6.8 | 230,767 | 7.8 | 579.8 | <0.001 |
| **Severe Sepsis (With Shock)** | 103,826 | 4.0 | 9,415 | 2.6 | 113,241 | 3.8 | 230767 | <0.001 |
| **CPAP use** | 118,393 | 4.6 | 41,305 | 11.4 | 159,698 | 5.4 | 28874.7 | <0.001 |

\*Student's *t*-test

### Death During Hospitalization

Patients with OSA had significantly lower odds of death (crude OR: 0.55, 95% CI [0.54, 0.56]), which remained significant following adjustment for multiple confounders (aOR: 0.72, 95% CI [0.70, 0.73]), and was similarly observed across all subgroups except UA (**Table 2**). The reduced odds of death of OSA patients were similar through years 2016 – 2019 (**Figure 1**).

**Table 2.**
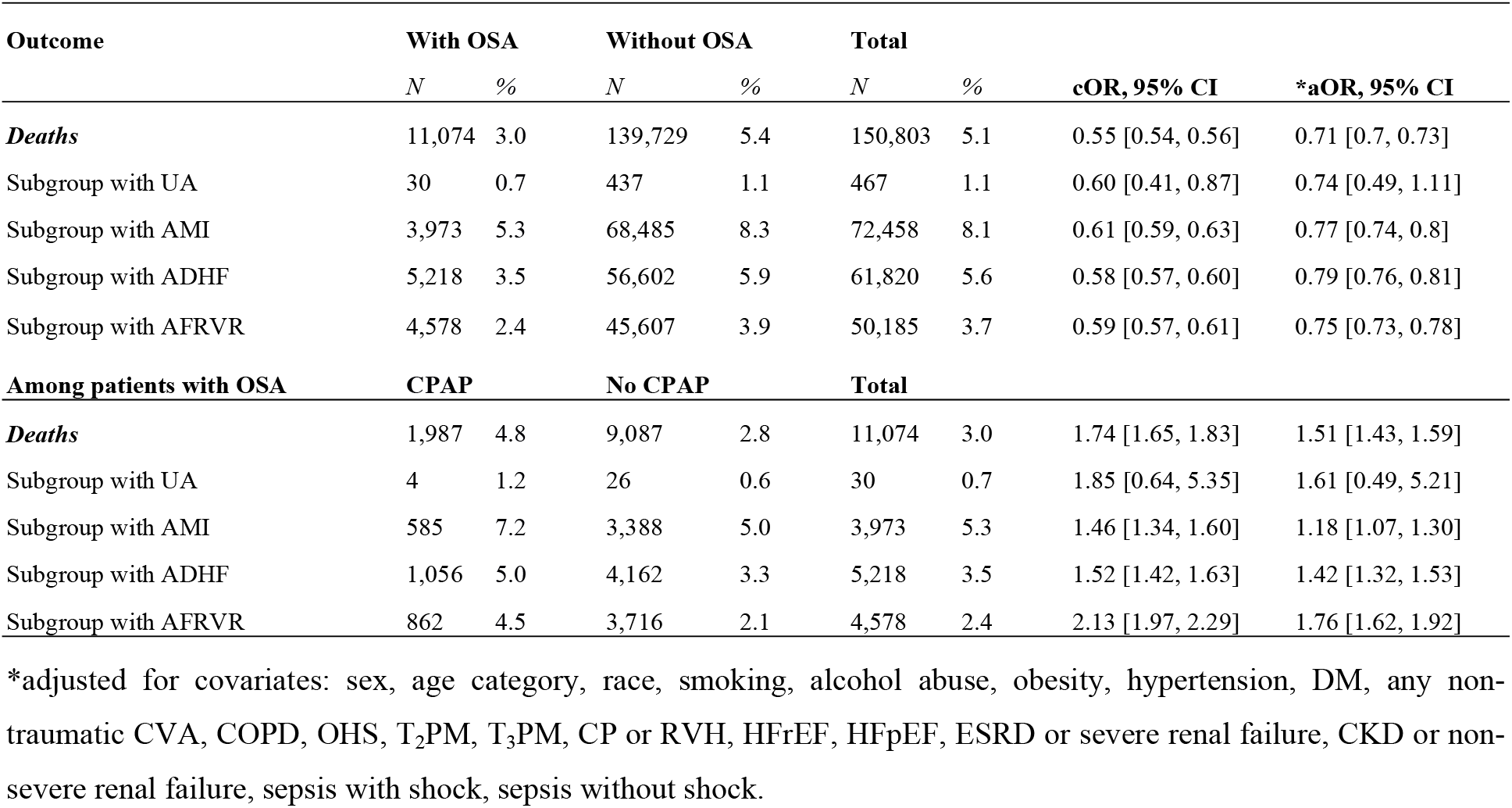
Outcomes for patient subgroups with Unstable Angina (UA), Acute Myocardial Infarction (AMI), Acute Decompensated Heart Failure (ADHF), and Atrial Fibrillation with Rapid Ventricular Respnse (AFRVR).

**Figure 1.**
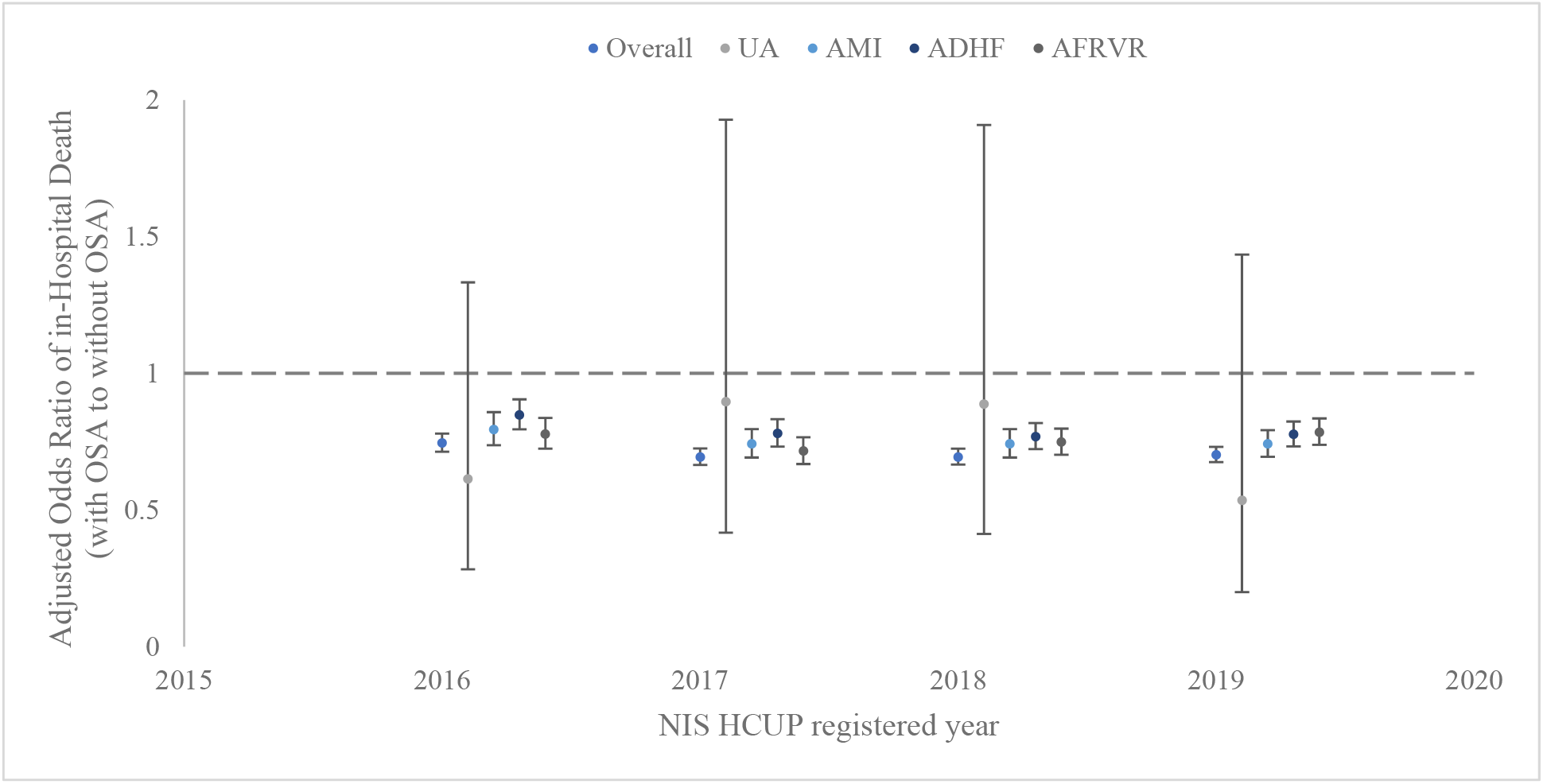
The reduced odds of in-hospital death when diagnosed with OSA through years 2016-2019.

Among patients with OSA, CPAP treatment during hospitalization was found to significantly increase the odds of death (crude OR: 1.74, 95% CI [1.65, 1.83]), which remained significant across all subgroups except UA, and following adjustment for multiple confounders (aOR: 1.51, 95% CI [1.44, 1.60]) (**Table 2**).

### Length of Stay and Total Hospital Charges

There were no statistically significant differences in length of stay (LOS) between patients with and without OSA (*p* = 0.60), and the difference was small following adjustment for confounders (adjusted mean difference: −0.06 days, 95% CI [−0.08, −0.03], *p* < 0.001) (**Table 3**). On the contrary, patients with OSA were charged significantly lower costs (adjusted mean difference: −$4709 95% CI [−5085, −4332]; *p* < 0.001), which was similar across subgroups except UA.

**Table 3.** Outcomes for patient subgroups with Unstable Angina (UA), Acute Myocardial Infarction (AMI), Acute Decompensated Heart Failure (ADHF), and Atrial Fibrillation with Rapid Ventricular Response

| Outcome | With OSA | Without OSA | Total | MD | F | p | †aMD, 95% CI | SE | *p |
| --- | --- | --- | --- | --- | --- | --- | --- | --- | --- |
| **Length of Stay (days)** | 6.06 (6.37) | 6.07 (7.47) | 6.07 (7.34) | -0.01 | 0.28 | 0.6 | -0.06 [-0.08, -0.03] | 0.01 | <0.001 |
| Subgroup with UA | 3.76 (4.16) | 3.56 (5.25) | 3.58 (5.15) | 0.20 | 5.90 | 0.015 | 0.04 [-0.13, 0.2] | 0.08 | 0.66 |
| Subgroup with AMI | 6.03 (6.38) | 5.78 (7.41) | 5.80 (7.32) | 0.25 | 82.76 | <0.001 | 0.06 [0.01, 0.11] | 0.03 | 0.03 |
| Subgroup with ADHF | 6.87 (6.91) | 6.99 (8.51) | 6.97 (8.31) | -0.12 | 25.87 | <0.001 | -0.01 [-0.06, 0.03] | 0.02 | 0.61 |
| Subgroup with AFRVR | 5.89 (6.19) | 6.1 (6.86) | 6.07 (6.77) | -0.21 | 162.93 | <0.001 | -0.34 [-0.38, -0.31] | 0.02 | <0.001 |
| **Total Charges (USD)** | 78,271 (113,969) | 83,495 (141,679) | 82,854 (138,590) | -5,224 | 449.05 | 0.032 | -4709<sup>§</sup> [-5085, -4332] | 192 | <0.001 |
| Subgroup with UA | 54,629 (66,933) | 51,959 (79,176) | 52,229 (78,029) | 2,671 | 4.62 | 0.007 | 560<sup>§</sup> [-1561, 2680] | 1082 | 0.605 |
| Subgroup with AMI | 100,616 (119,978) | 102,231 (145,519) | 102,095 (143,551) | -1,615 | 8.65 | 0.003 | -5408<sup>§</sup> [-6250, -4567] | 429 | <0.001 |
| Subgroup with ADHF | 82,805 (135,474) | 90,285 (178,666) | 89,294 (173,582) | -7,480 | 235.13 | <0.001 | -4344<sup>§</sup> [-5061, -3627] | 366 | <0.001 |
| Subgroup with AFRVR | 73,314 (104,146) | 75,345 (114,786) | 75,057 (113,341) | -2,031 | 52.49 | <0.001 | -976<sup>§</sup> [-1417, -534] | 225 | <0.001 |
| **Among patients with OSA** | **used CPAP** | **did not use CPAP** | **Total** | **MD** | **F** | **p** | **aMD, 95% CI** | **SE** | **\*p** |
| **Length of Stay (days)** | 7.78 (7.43) | 5.84 (6.19) | 6.06 (6.37) | 1.94 | 3395.9 | <0.001 | 1.49<sup>†</sup> [1.43, 1.55] | 0.03 | <0.001 |
| **Total Charges (USD)** | 96,909 (125,732) | 75,887 (112,152) | 78,271 (113,969) | 21,023 | 1236.2 | <0.001 | 1168<sup>§</sup> [273, 2062] | 456 | 0.011 |

Numbers in parentheses are standard deviations of respective means
MD = mean difference
aMD = adjusted mean difference
\*significance level for one-tailed test, adjusted for multiple comparisons (Bonferroni method)

Patients with OSA who were treated with CPAP during their hospitalization spent significantly more inpatient days (adjusted mean difference: 1.49 days, 95% CI [1.43, 1.55]; *p* < 0.001), and were charged significantly higher costs (adjusted mean difference: $1168, 95% CI [273, 2,062]; *p* = 0.011).

## Discussion

In our study of about 3 million hospitalization records, patients with OSA were found to have significantly lower in-hospital mortality. Additionally, there was no significant difference in LOS between patients with and without OSA, yet patients with OSA were found to have lower hospital charges. OSA patients treated with CPAP were at increased odds of in-hospital death, had longer stay duration and were charged larger hospital costs.

Some studies have reported significantly lower in-hospital mortality in AMI patients with OSA not treated with CPAP during index hospitalization^1,7^. Our study included a larger selection of cardiovascular disease, while aiming to capture the acuity of the encounter: AMI, ADHF, and AFRVR; and our results were consistent with the findings of these studies^1,7^. Our results also showed that patients with OSA were more likely to be younger, male, smokers, obese, and have stroke, COPD, renal failure, and heart failure, which was in line with findings of many studies reporting associations between cardiovascular disease and OSA^8–12^ (**Table 1**).

While acknowledging the myriad reported negative cardiovascular outcomes associated with OSA^13–18^, we postulate that the observed survival benefit may be due to ischemic preconditioning^19,20^, a potential cardioprotective phenomenon caused by OSA-induced chronic intermittent hypoxia^2^. Intermittent episodes of hypoxia result in miniature ischemic episodes that confer adaptation and protection from future infarctions and life-threatening arrhythmias^21–24^. Our findings also showed that patients with OSA who were hospitalized with acute cardiovascular disease and were treated with CPAP had higher mortality. Contrary to expectations that aggressively treating hospitalized OSA patients with CPAP would dramatically improve outcomes^25^, we postulate that OSA patients might be chronically dependent on intermittent hypoxia, and the routine use of CPAP during the acute cardiovascular encounter (e.g. AMI, ADHF, AFRVR) could neutralize the potential benefits of ischemic preconditioning, much similar to how routine administration of oxygen to normoxic patients with AMI confers no additional benefit and may even cause harm^26,27^.

Although our results are thought-provoking, there are several notable limitations to our analysis. First, we used ICD-10-CM codes to identify OSA patients, thereby lacking data on disease severity, as well as the use of and compliance with CPAP treatment prior to hospitalization, and thereby increasing the risk for selection bias. Second, we did not account for many important diagnostic and therapeutic procedures performed during hospitalizations, such as thrombolysis, diagnostic angiography, PCI, and CABG, all of which can impact mortality, length of stay and hospital charges^7^. Last, although we applied multivariable adjustment for demographic and clinical confounders, there is possibility of unaccounted for confounding variables. Notwithstanding these limitations, our results were consistent with previous research^2,7,23,28^.

## Conclusion

Patients with recognized OSA who are hospitalized with AMI, ADHF and AF have significantly lower in-hospital mortality. However, OSA patients treated with CPAP had increased in-hospital mortality, resource utilization and length of stay. Randomized controlled trials focusing on severity of sleep apnea in different high-risk patient groups are needed to clarify the current practice of initiating CPAP in the acute setting, much like how the old practice of routine oxygen supplementation in acute myocardial infarction was revisited.

## Data Availability

The authors confirm that the data supporting the findings of this study are available within the article [and/or] its supplementary materials.

## Supplemental Material

**Supplemental Table 1.** Specific ICD-10-CM, ICD-10-PCS, and AHRQ codes used to identify conditions of interest

| Diagnosis, Condition, Procedure | Query Code | Code Type |
| --- | --- | --- |
| Unstable Angina (UA) | I20 | ICD-10-CM |
| Acute Myocardial Infarction (AMI) | I21, I22 | ICD-10-CM |
| Acute Decompensated Heart Failure (ADHF) | I5021, I5023, I5031, I5033, I5041, I5043, I50811, I50813, I5084 | ICD-10-CM |
| Atrial Fibrillation with Rapid Ventricular Response (AFRVR) | I480 | ICD-10-CM |
| Obstructive Sleep Apnea (OSA) | G4733 | ICD-10-CM |
| Central Sleep Apnea (CSA) | G4739 | ICD-10-CM |
| Snoring | R0683 | ICD-10-CM |
| Obesity | E66 | ICD-10-CM |
| Overweight | E663 | ICD-10-CM |
| Tobacco Use | F1721, Z720, Z87891 | ICD-10-CM |
| Alcohol Abuse | ALCOHOL | AHRQ |
| Obesity Hypoventilation Syndrome (OHS) | E662 | ICD-10-CM |
| Heart Failure with Reduced Ejection Fraction (HFrEF) | I502 | ICD-10-CM |
| Heart Failure with Preserved Ejection Fraction (HFpEF) | I503 | ICD-10-CM |
| Sepsis | A40, A41 | ICD-10-CM |
| Septic Shock | R6521 | ICD-10-CM |
| Non-Traumatic Cerebrovascular Accidents (CVA) | I60, I61, I62, I63 | ICD-10-CM |
| Chronic Obstructive Pulmonary Disorder (COPD) | J42, J43, J44 | ICD-10-CM |
| Pulmonary Hypertension Type II | I2772 | ICD-10-CM |
| Pulmonary Hypertension Type III | I2773 | ICD-10-CM |
| Right Ventricular Hypertrophy (RVH) | I279 | ICD-10-CM |
| Cor Pulmonale | I2781 | ICD-10-CM |
| Acute Kidney Injury (AKI) | N17 | ICD-10-CM |
| Chronic Kidney Disease (CKD) | N18 | ICD-10-CM |
| Kidney Failure, non-specified | N19 | ICD-10-CM |
| End-Stage Renal Disease (ESRD) | N186 | ICD-10-CM |
| Severe Renal Failure | RENLFL_SEV | AHRQ |
| Diabetes Mellitus (DM) | DIAB_UX, DIAB_CX | AHRQ |
| Hypertension | HTN_UX, HTN_CX | AHRQ |
| Continuous Positive Airway Pressure (CPAP) | 5A09357, 5A09457, 5A09557 | ICD-10-PCS |

## References

1. Yeghiazarians Y, Jneid H, Tietjens JR, et al. Obstructive Sleep Apnea and Cardiovascular Disease: A Scientific Statement From the American Heart Association. Circulation 2021;144(3):e56–e67. (In eng). DOI: 10.1161/cir.0000000000000988.

2. Shah N, Redline S, Yaggi HK, et al. Obstructive sleep apnea and acute myocardial infarction severity: ischemic preconditioning? Sleep Breath 2013;17(2):819–26. (In eng). DOI: 10.1007/s11325-012-0770-7.

3. McEvoy RD, Antic NA, Heeley E, et al. CPAP for Prevention of Cardiovascular Events in Obstructive Sleep Apnea. N Engl J Med 2016;375(10):919–31. (In eng). DOI: 10.1056/NEJMoa1606599.

4. Lyons PG, Zadravecz FJ, Edelson DP, Mokhlesi B, Churpek MM. Obstructive sleep apnea and adverse outcomes in surgical and nonsurgical patients on the wards. J Hosp Med 2015;10(9):592–8. (In eng). DOI: 10.1002/jhm.2404.

5. Healthcare Cost and Utilization Project (HCUP) Statistical Briefs. Rockville (MD): Agency for Healthcare Research and Quality (US); 2006.

6. Steiner C, Elixhauser A, Schnaier J. The healthcare cost and utilization project: an overview. Eff Clin Pract 2002;5(3):143–51. (In eng).

7. Mohananey D, Villablanca PA, Gupta T, et al. Recognized Obstructive Sleep Apnea is Associated With Improved In-Hospital Outcomes After ST Elevation Myocardial Infarction. J Am Heart Assoc.

8. Schäfer H, Koehler U, Ewig S, Hasper E, Tasci S, Lüderitz B. Obstructive sleep apnea as a risk marker in coronary artery disease. Cardiology 1999;92(2):79–84. (In eng). DOI: 10.1159/000006952.

9. Prisco DL, Sica AL, Talwar A, et al. Correlation of pulmonary hypertension severity with metrics of comorbid sleep-disordered breathing. Sleep Breath 2011;15(4):633–9. (In eng). DOI: 10.1007/s11325-010-0411-y.

10. Nieto FJ, Young TB, Lind BK, et al. Association of sleep-disordered breathing, sleep apnea, and hypertension in a large community-based study. Sleep Heart Health Study. Jama 2000;283(14):1829–36. (In eng). DOI: 10.1001/jama.283.14.1829.

11. Koskenvuo M, Kaprio J, Telakivi T, Partinen M, Heikkilä K, Sarna S. Snoring as a risk factor for ischaemic heart disease and stroke in men. Br Med J (Clin Res Ed) 1987;294(6563):16–9. (In eng). DOI: 10.1136/bmj.294.6563.16.

12. Peker Y, Kraiczi H, Hedner J, Löth S, Johansson A, Bende M. An independent association between obstructive sleep apnoea and coronary artery disease. Eur Respir J 1999;14(1):179–84. (In eng). DOI: 10.1034/j.1399-3003.1999.14a30.x.

13. Wang LJ, Pan LN, Yan RY, Quan WW, Xu ZH. Obstructive sleep apnea increases heart rhythm disorders and worsens subsequent outcomes in elderly patients with subacute myocardial infarction. J Geriatr Cardiol 2021;18(1):30–38. (In eng). DOI: 10.11909/j.issn.1671-5411.2021.01.002.

14. Kanagala R, Murali NS, Friedman PA, et al. Obstructive sleep apnea and the recurrence of atrial fibrillation. Circulation 2003;107(20):2589–94. (In eng). DOI: 10.1161/01.Cir.0000068337.25994.21.

15. Yumino D, Tsurumi Y, Takagi A, Suzuki K, Kasanuki H. Impact of obstructive sleep apnea on clinical and angiographic outcomes following percutaneous coronary intervention in patients with acute coronary syndrome. Am J Cardiol 2007;99(1):26–30. (In eng). DOI: 10.1016/j.amjcard.2006.07.055.

16. Uchôa CHG, Pedrosa RP, Javaheri S, et al. OSA and Prognosis After Acute Cardiogenic Pulmonary Edema: The OSA-CARE Study. Chest 2017;152(6):1230–1238. (In eng). DOI: 10.1016/j.chest.2017.08.003.

17. Wang H, Parker JD, Newton GE, et al. Influence of obstructive sleep apnea on mortality in patients with heart failure. J Am Coll Cardiol 2007;49(15):1625–1631. (In eng). DOI: 10.1016/j.jacc.2006.12.046.

18. Khayat R, Jarjoura D, Porter K, et al. Sleep disordered breathing and post-discharge mortality in patients with acute heart failure. Eur Heart J 2015;36(23):1463–9. (In eng). DOI: 10.1093/eurheartj/ehu522.

19. Tomai F, Crea F, Chiariello L, Gioffrè PA. Ischemic preconditioning in humans: models, mediators, and clinical relevance. Circulation 1999;100(5):559–63. (In eng). DOI: 10.1161/01.cir.100.5.559.

20. Stokfisz K, Ledakowicz-Polak A, Zagorski M, Zielinska M. Ischaemic preconditioning - Current knowledge and potential future applications after 30 years of experience. Adv Med Sci 2017;62(2):307–316. (In eng). DOI: 10.1016/j.advms.2016.11.006.

21. Sforza E, Roche F. Chronic intermittent hypoxia and obstructive sleep apnea: an experimental and clinical approach. Hypoxia (Auckl) 2016;4:99–108. (In eng). DOI: 10.2147/hp.S103091.

22. Ferdinandy P, Schulz R, Baxter GF. Interaction of cardiovascular risk factors with myocardial ischemia/reperfusion injury, preconditioning, and postconditioning. Pharmacol Rev 2007;59(4):418–58. (In eng). DOI: 10.1124/pr.107.06002.

23. Ozeke O, Ozer C, Gungor M, Celenk MK, Dincer H, Ilicin G. Chronic intermittent hypoxia caused by obstructive sleep apnea may play an important role in explaining the morbidity-mortality paradox of obesity. Med Hypotheses 2011;76(1):61–3. (In eng). DOI: 10.1016/j.mehy.2010.08.030.

24. Neckár J, Ostádal B, Kolár F. Myocardial infarct size-limiting effect of chronic hypoxia persists for five weeks of normoxic recovery. Physiol Res 2004;53(6):621–8. (In eng).

25. De Torres-Alba F, Gemma D, Armada-Romero E, Rey-Blas JR, López-de-Sá E, López-Sendon JL. Obstructive sleep apnea and coronary artery disease: from pathophysiology to clinical implications. Pulm Med 2013;2013:768064. (In eng). DOI: 10.1155/2013/768064.

26. Hofmann R, Befekadu Abebe T, Herlitz J, et al. Routine Oxygen Therapy Does Not Improve Health-Related Quality of Life in Patients With Acute Myocardial Infarction-Insights From the Randomized DETO2X-AMI Trial. Front Cardiovasc Med 2021;8:638829. (In eng). DOI: 10.3389/fcvm.2021.638829.

27. Hofmann R, James SK, Jernberg T, et al. Oxygen Therapy in Suspected Acute Myocardial Infarction. N Engl J Med 2017;377(13):1240–1249. (In eng). DOI: 10.1056/NEJMoa1706222.

28. Lavie L, Lavie P. Reduced Cardiovascular Morbidity in Obesity-Hypoventilation Syndrome: An Ischemic Preconditioning Protective Effect? Chest 2016;150(1):5–6. (In eng). DOI: 10.1016/j.chest.2016.02.659.

